## supplement files for "ACVR2A Facilitates Trophoblast Cell Invasion through TCF7/c-JUN Pathway in Pre-eclampsia Progression"

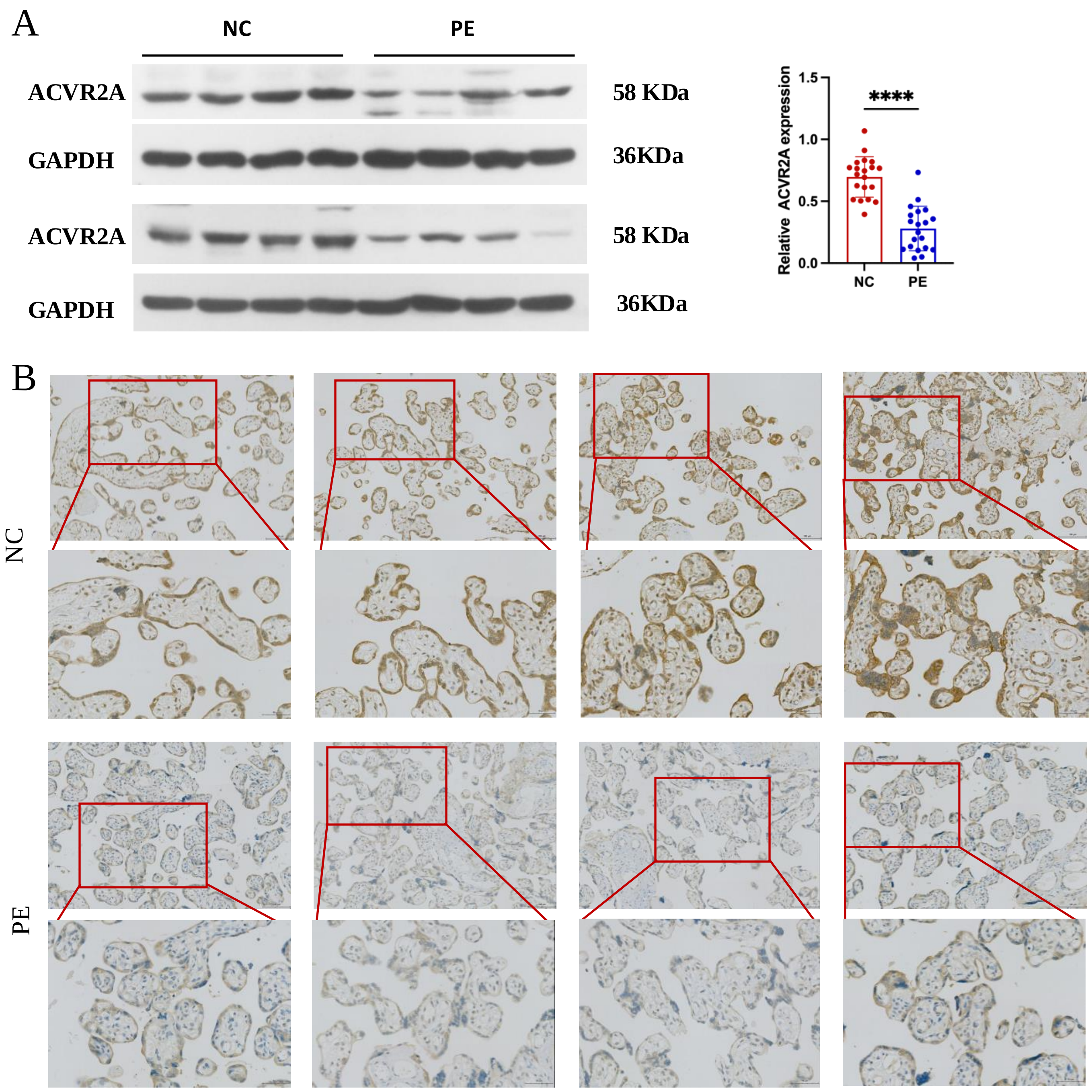


Figure S1. Downregulation of ACVR2A Expression in Preeclampsia Placental Tissue: Insights from Western Blot and Immunohistochemistry.

(A) Western blot analysis demonstrated reduced levels of ACVR2A in preeclampsia placental tissue (n = 20) compared to control placentas.

(B) Immunohistochemical staining using rabbit IgG anti-human ACVR2A antibody on sections from normal control pregnant women and preeclampsia placentas. Sections were counterstained with hematoxylin. ACVR2A levels were markedly lower in patients with preeclampsia (n = 10) compared to normal control subjects.

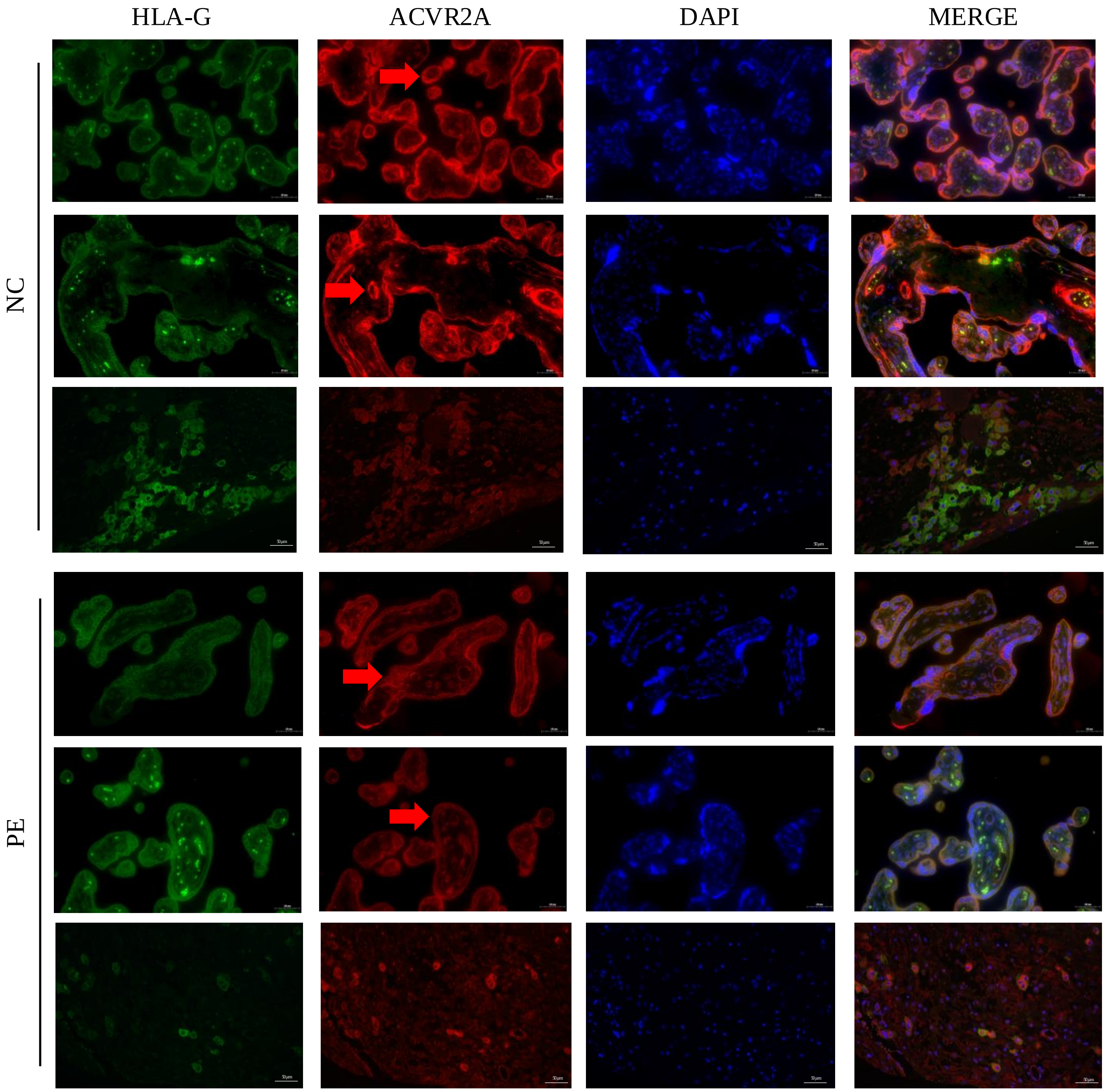


Figure S2. Reduced ACVR2A Expression in Trophoblast Cells of Preeclampsia Placental Tissue Revealed by Immunofluorescence Co-localization with HLA-G.Immunofluorescence co-localization of rabbit IgG anti-human ACVR2A antibodies and HLA-G antibodies (a marker of extrachorionic trophoblastic cells) in normal control and preeclampsia maternal placenta. The expression pattern of the ACVR2A antibody closely resembles that of the HLA-G antibody, primarily expressed in EVT cells.

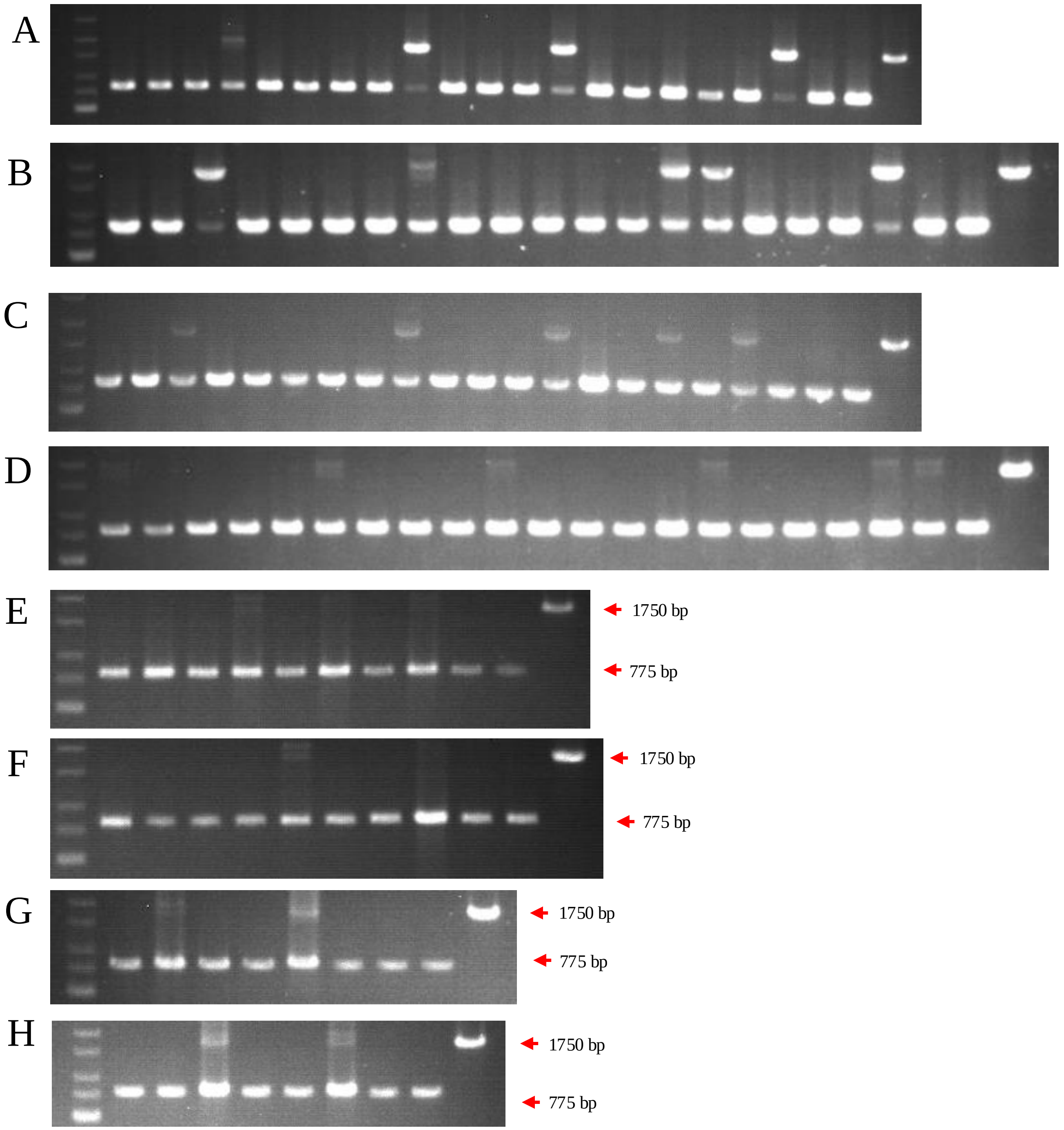


Figure S3. Genotype identification led to the selection of three pairs of HTR8/SVneo and JAR cell lines with complete ACVR2A knockout.
(A) Identification of the first batch of ACVR2A gene knockout monoclonal cell lines in the HTR8/SVneo cell line.
(B) Identification of the first batch of ACVR2A gene knockout monoclonal cell lines in the JAR cell line.
(C) Identification of the second batch of ACVR2A gene knockout monoclonal cell lines in the HTR8/SVneo cell line.
(D) Identification of the second batch of ACVR2A gene knockout monoclonal cell lines in the JAR cell line.
(E) Identification of the third batch of ACVR2A gene knockout monoclonal cell lines in the HTR8/SVneo cell line.
(F) Identification of the third batch of ACVR2A gene knockout monoclonal cell lines in the JAR cell line.
(G) Identification of the fourth batch of ACVR2A gene knockout monoclonal cell lines in the HTR8/SVneo cell line.
(H) Identification of the fourth batch of ACVR2A gene knockout monoclonal cell lines in the JAR cell line.

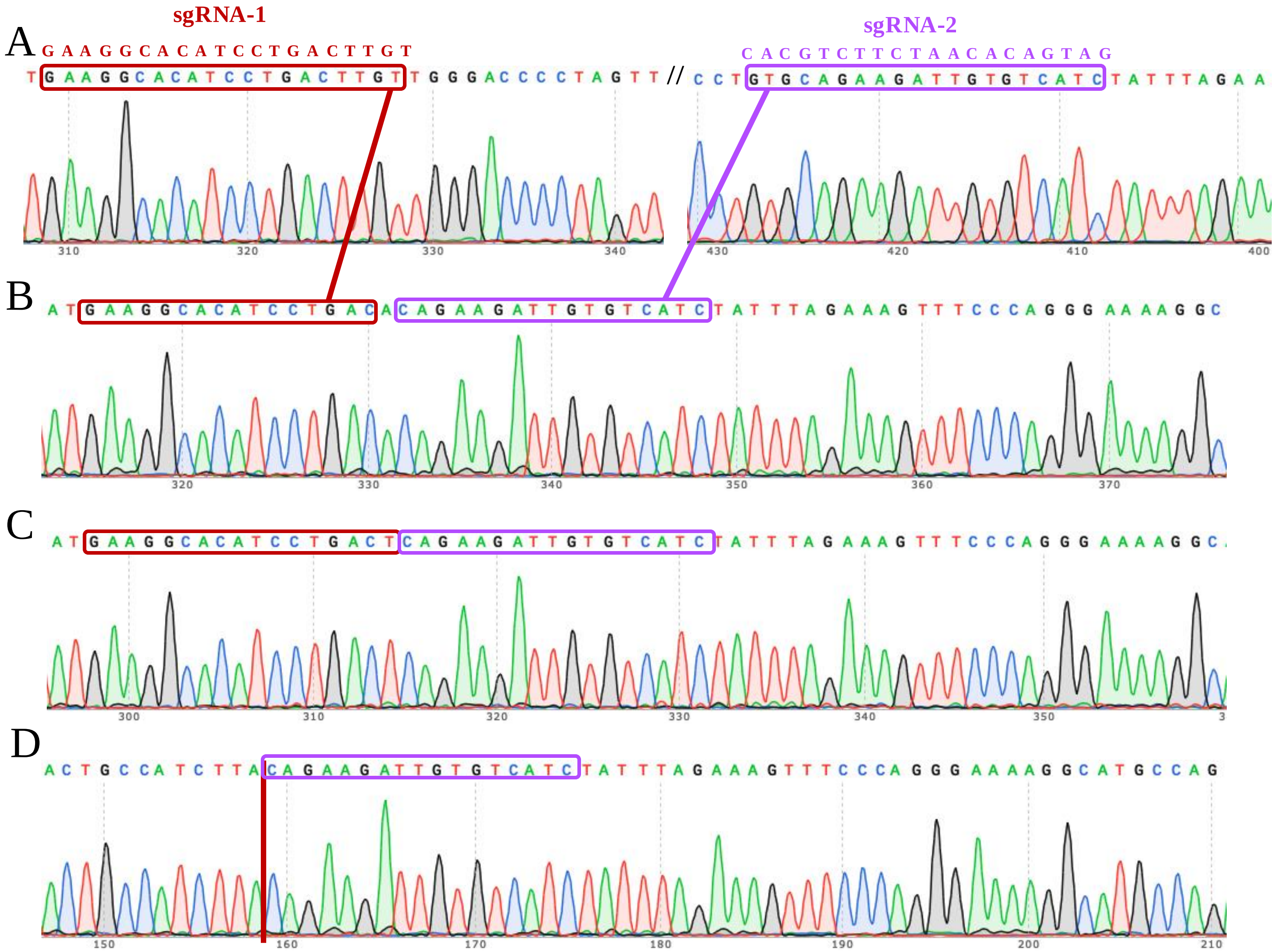


Figure S4. The complete knockout of the ACVR2A gene in HTR8/Svneo monoclonal cell lines was confirmed through Sanger sequencing electrophoresis.

The Sanger sequencing technique was employed to validate the successful knockout (KO) of the ACVR2A gene in these specific trophoblast cell lines. Each lane corresponds to a distinct monoclonal cell line: HTR8/SVneo wild type（A）, HTR8/SVneo KO-1（B), HTR8/SVneo KO-2(C), and HTR8/SVneo KO-3(D). The electropherograms provide a visual representation of the DNA fragments, confirming the precise and targeted deletion of the ACVR2A gene in the knockout cell lines. The distinct peaks and patterns in the electropherograms indicate the specific alterations made during the gene editing process. These sequencing results validate the efficacy of CRISPR/Cas9-mediated ACVR2A gene knockout in the HTR8/Svneo trophoblast cell lines.

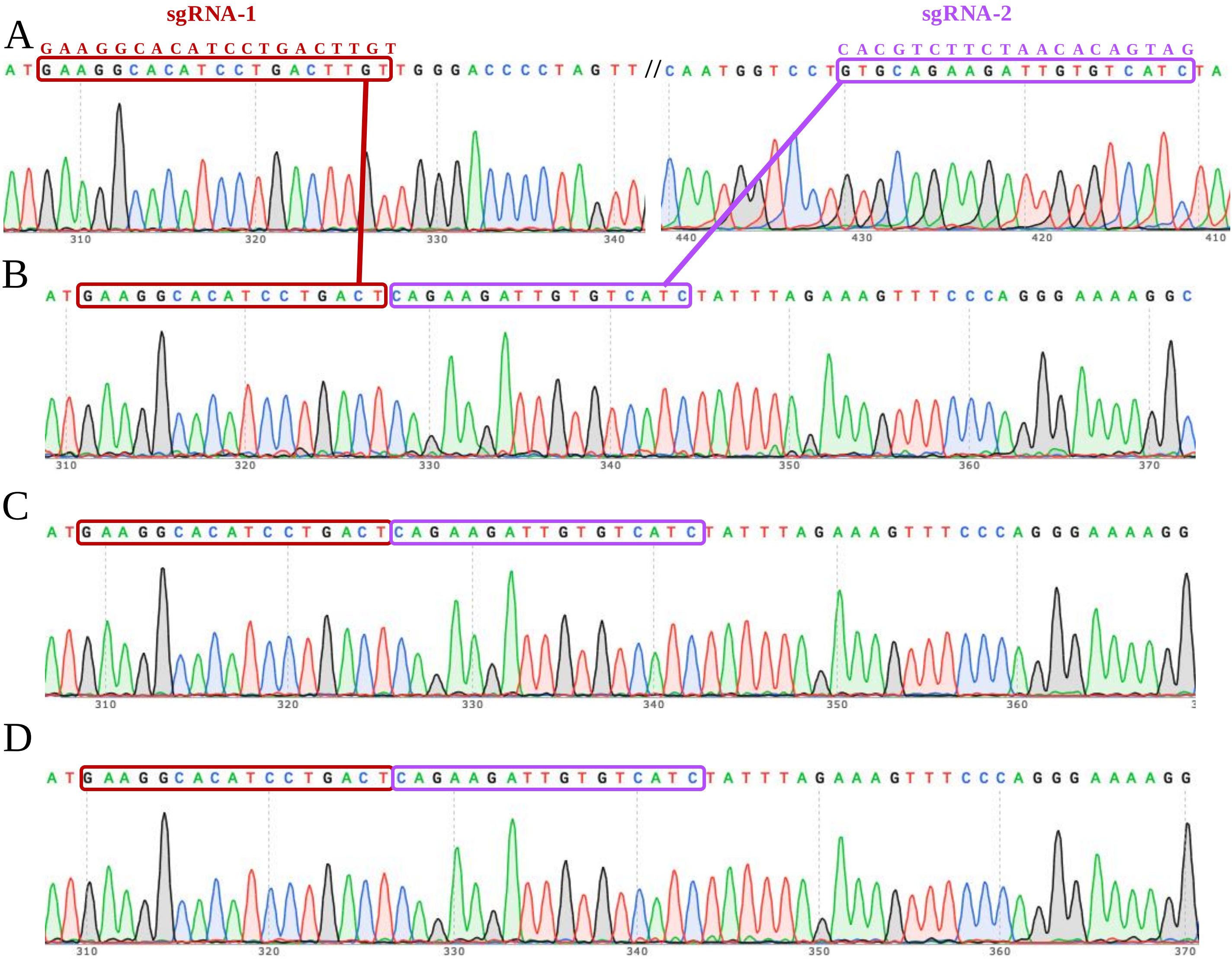


Figure S5. The complete knockout of the ACVR2A gene in JAR monoclonal cell lines was confirmed through Sanger sequencing electrophoresis.

Each lane corresponds to a specific monoclonal cell line: JAR wild type(A), JAR KO-1(B), JAR KO-2(C), and JAR KO-3(D). The electropherograms capture the unique DNA fragment patterns generated during the gene editing process, confirming the successful deletion of the ACVR2A gene in the knockout cell lines. The distinctive peaks and sequences in the electropherograms offer a visual validation of the precision and accuracy of the CRISPR/Cas9 gene editing technique in inducing targeted genetic changes. These sequencing results serve as a crucial verification step, establishing the reliability of the JAR-ACVR2A-KO monoclonal cell lines for subsequent investigations into the functional implications of ACVR2A in trophoblast cells and its potential relevance to conditions such as preeclampsia.

#### Table S1 Clinical characteristics of pre-eclamptic and normal control pregnancies.

| Variable | PE group (n=20, mean ± SD ) | NC group (n=20, mean ± SD) |
|---|---|---|
| Age [a] | 33 ± 3 | 31.4 ± 2.44 |
| Predelivery BMI [a] (kg/m2) | 24.79± 3.22 | 23.34 ± 3.39 |
| Systolic blood pressure [a] (mmHg) | 149.50 ± 7.31 | 113.79 ± 9.97 |
| Diastolic blood pressure [a] (mmHg) | 97.65 ± 6.92 | 72.63 ± 5.88 |
| Gestational age [a] (week) | 36 (34, 38) | 38 (37, 39) |
| Neonatal Birth Weight (g) | 2738.75 ± 694.96 | 3221.50 ± 242.75 |

Continuous variables that conform to the normal distribution are represented by the mean sigma standard deviation, and those that do not conform to the normal distribution are represented by the median (P25, P75)
a The Mann-Whitney test was used for statistical analysis, median (IQR).
b The Chi-square test was used for statistical analysis.

#### Table S2. Sequences of the primers for RT-qPCR.

| Primer | Sequence |
|---|---|
| GAPDH-F | 5’- AAAAGCATCACCCGGAGGAGAA -3’ |
| GAPDH-R | 5’- GATAACCTGGCTTCTGCGTCGT -3’ |
| C-JUN-F | 5’- GATAACCTGGCTTCTGCGTCGT -3’ |
| C-JUN-R | 5’- TGCTGCGTTAGCATGAGTTGGC -3’ |
| TCF7-F | 5’- CTGACCTCTCTGGCTTCTACTC -3’ |
| TCF7-R | 5’- CAGAACCTAGCATCAAGGATGGG -3’ |
| TCF7L1-F | 5’- TCAAGGACACGAGGTCACCATC -3’ |
| TCF7L1-R | 5’- GGAGAAGTGGTCATTGCTGTAGG -3’ |
| Wnt3-F | 5’- GCGTGTTAGTGTCCAGGGAGTT -3’ |
| Wnt3-R | 5’- TGAGGTGCATGTGGTCCAGGAT -3’ |
| Wnt4-F | 5’- ATGAACCTCCACAACAATGAG -3’ |
| Wnt4-R | 5’- ACCATCAAACTTCTCCTTCAG -3’ |
| SMAD4-F | 5’- CTACCAGCACTGCCAACTTTCC -3’ |
| SMAD4-R | 5’- CCTGATGCTATCTGCAACAGTCC -3’ |
| CCND1-F | 5’- TCTACACCGACAACTCCATCCG -3’ |
| CCND1-R | 5’- TCTGGCATTTTGGAGAGGAAGTG -3’ |
| ACVR2A-F | 5’- GCCAGCATCCATCTCTTGAAGAC -3’ |
| ACVR2A-R | 5’- GATAACCTGGCTTCTGCGTCGT -3’ |

#### Table S3. Sequences of the genotyping primers.

| Primer | Sequence |
|---|---|
| ACVR2A-F | 5’- ACTGATACTGCTCAGTGGTGAC -3’ |
| ACVR2A-R | 5’- CCCTTGTTCATAACCCAGGTC -3’ |

### Table S4. Details of immunohistochemical antibodies

| Antibody | corporation | Cat | concentration |
| --- | --- | --- | --- |
| ACVR2A | Thermo | PA5-95374 | 1:300 |
| HLA-G | proteintech | 66447-1-IG | 1:100 |
| Wnt3 | bioss | bs-1700R | 1:100 |
| Wnt4 | bioss | bs-20786R | 1:100 |
| c-JUN | Servicebio | GB11071 | 1:1000 |
| CCND1 | Servicebio | GB111372 | 1:600 |
| TCF7L1 | bioss | bs-12891R | 1:100 |
| TCF7L2 | bioss | bs-1280R | 1:100 |
| SMAD4 | bioss | bsm-52225R | 1:100 |
| SMAD1/5 | bioss | bs-2973R | 1:100 |
| pSMAD1/5/9 | CellSignalingTechnology | 13820T | 1:100 |
